# Nurse-Led Heart Failure Education and Patient Outcomes: A Mixed-Methods Prospective Registry Study

**DOI:** 10.64898/2026.11.08.26367583

**Authors:** Erika Nagata, Reona Sasamoto, Ami Endo, Shino Miura, Haruna Goto, Hiroaki Sukegawa, Yoshinori Mano, Takahiro Ohki, Mitsuaki Sawano

## Abstract

**Background:** Patient understanding of heart failure (HF) self-care varies widely and is inconsistently associated with clinical outcomes. We examined knowledge acquisition and patient experiences related to structured HF education using a patient handbook.

**Methods and Results:** Between April and December 2021, 56 patients hospitalized for acute HF (mean age 74 years, 35% women, 46% HFrEF) were enrolled from 176 screened patients at a single center. A 20-item knowledge assessment was administered before and after nurse led HF education and at first outpatient follow-up. Thirty patients met criteria for knowledge improvement (≥5-point increase or baseline ≥15), while 25 did not. Knowledge gaps were most evident in exercise, medication effects, salt restriction, and recognition of exacerbating factors. Patients with knowledge improvement had higher post-education scores (16 vs. 12), sustained knowledge at follow-up, higher serum albumin levels, better inpatient dietary intake. One-year mortality and HF readmission did not differ. A nested qualitative analysis of four readmitted patients identified 94 codes grouped into 9 categories, highlighting personalized symptom recording, persistent self-management challenges despite handbook use, enhanced communication with healthcare professionals, reflection on future care, and a desire for earlier HF education.

**Conclusions:** Patients who achieved higher HF knowledge scores demonstrated better nutritional status but similar one-year clinical outcomes. Qualitative findings underscore ongoing challenges in long-term HF self-care.

## Introduction

Heart failure (HF) represents a growing global health burden, particularly in aging societies such as Japan.^1,2^ Despite advances in pharmacological and device-based therapies, rehospitalization and mortality rates remain high. Updated guidelines emphasize patient education and multidisciplinary self-care support to reduce HF hospitalizations and mortality, with attention to symptom monitoring and factors like frailty and psychosocial factors.^3^ To strengthen patient support, the Japanese Circulation Society (JCS) launched the Certified Heart Failure Educator program in 2021, and admission to facilities with such educators has been associated with improved process measures and lower in-hospital mortality, underscoring the value of structured, expert-led education in routine practice.^4^

Among certified heart failure educators, registered nurses form the foundation of patient care and communication. Given their continuous involvement in both inpatient and outpatient management, registered nurses are uniquely positioned to deliver education and reinforce self-care behaviors. Consequently, nurse-led self-care education has attracted considerable attention, as nurses are often central to ongoing patient support and disease management.^5^ Further, a meta-analysis reported that nurse-led education significantly reduced the risks of readmission and adverse outcomes in patients with HF, highlighting its potential clinical value.^6^ However, improvements in quality of life and disease knowledge have not been consistently observed, and the specific knowledge domains in which educational efforts may be less effective remain poorly defined. Moreover, the ways in which patients’ beliefs, understanding, and lived experiences of education relate to knowledge acquisition and clinical outcomes are not well characterized, with factors such as nutrition, cognition, social support, and engagement likely contributing to variability in educational effectiveness.

To address these gaps, we conducted a single-center, prospective mixed-methods registry of patients hospitalized with acute heart failure. By integrating quantitative assessments of knowledge, quality of life, and clinical characteristics with qualitative patient interviews, this study aimed to clarify how heart failure knowledge varies across individuals, to identify specific domains in which educational understanding differs, and to explore patient-reported experiences that may contextualize these differences. Through this approach, we sought to better characterize factors associated with variation in educational uptake and to inform more effective, real-world strategies for heart failure education.

## Methods

### Study Design

This single-center, prospective mixed-methods heart failure registry was conducted in the Department of Cardiology at Tokyo Dental College Ichikawa General Hospital, a secondary care hospital in Ichikawa City, Chiba, Japan. The study was approved by the institutional Ethics Review Board (Approval No. I-19-96) and conducted in accordance with the Declaration of Helsinki. Written informed consent was obtained from all participants prior to enrollment.

Eligible patients were aged 20 years or older and admitted with a first episode of acute heart failure. Inclusion required New York Heart Association (NYHA) functional class II or higher symptoms (e.g., dyspnea, fatigue, or edema) plus a brain natriuretic peptide (BNP) level ≥100 pg/mL or N-terminal pro-BNP ≥400 pg/mL. Patients with acute coronary syndrome who developed heart failure during hospitalization were excluded. For the substudy cohort, additional exclusion criteria were to cognitive impairment or lack of self-care capacity precluding informed consent, as determined by two or more investigators due to the requirement for knowledge testing.

The registry included consecutive patients hospitalized for acute heart failure. Patients with sufficient cognitive function to participate in educational assessments and questionnaires comprised the main analytical cohort, while a separate qualitative cohort included patients who underwent semi-structured nurse-led interviews during or shortly after rehospitalization for heart failure. All patients received standard inpatient care, including nurse-led discharge education supported by a structured Heart Failure Handbook developed through a multidisciplinary process. Patients in the main cohort completed a 20-item heart failure knowledge assessment (**Supplemental Table 1**) before and after HF eduation during index hospitalization and again at the first outpatient follow-up. Similarily, the Japanese version of the KCCQ-12 was assessed prior to discharge and again at the first outpatient follow-up. This report presents findings from participants enrolled between April and December 2021 with one-year follow-up thereafter.

Among the patients who were readmitted for heart failure, interviews were conducted under the lead of a nurse investigator to ensure consistency and depth of inquiry (**Supplemental Table 2**). Using a semi-structured format of approximately 30 minutes, the interviews were guided by an interview tool designed to explore patients’ experiences of incorporating the Heart Failure Handbook into their daily lives. Participants were asked about their routines for using the handbook, the perceived benefits and challenges, and how discharge education shaped their self-management practices at home. Each interview was conducted face-to-face, recorded with consent, and transcribed verbatim. The data were analyzed using a qualitative descriptive approach, beginning with line-by-line coding of the transcripts, followed by grouping of codes into subcategories, and further abstraction into broader categories.

### Key Variables and Outcomes

#### Baseline characteristics

Data collected included baseline demographics, risk factors, comorbidities, medications, heart failure knowledge, quality of life, and clinical outcomes. Baseline clinical risk was assessed using the Meta-Analysis Global Group in Chronic Heart Failure (MAGGIC) risk score. The MAGGIC score is a validated prognostic tool derived from pooled datasets of over 39,000 HF patients worldwide that has also been validated in the Japanese heart failure population.^9,10^ It incorporates 13 clinical variables, including age, sex, systolic blood pressure, body mass index, left ventricular ejection fraction, NYHA class, serum creatinine, presence of diabetes, current smoking, chronic obstructive pulmonary disease, time since HF diagnosis, use of beta-blockers, and use of ACE inhibitors or ARBs. Scores typically range from 0 to over 50, with higher values indicating greater predicted mortality risk. Salt-reduced, standardized meals were served to all patients three times daily during hospitalization. Food consumption was assessed at nine meal points over the three days preceding discharge and averaged for analysis. In this study, the *main dish* refers to the staple component of hospital meals, which is generally richer in carbohydrates and accounts for approximately 70% of total energy intake, whereas *side dishes* refer to accompanying components that are generally richer in protein and minerals, contributing approximately 30% of total intake and supporting muscle maintenance and micronutrient balance.

#### Knowledge assessment

Knowledge improvement was defined as either a baseline score of ≥15 out of 20 on the original structured 20-item test or an increase of ≥5 points following nurse-led predischarge education using the Heart Failure Handbook. Each item was equally weighted, yielding a maximum score of 20. Patients meeting these criteria were classified as the knowledge-improved group (Group A), whereas those who did not were classified as the non-improved group (Group B). We further examined: (1) items demonstrating a ≥40% difference in correct response rates between groups; (2) items with high correct response rates in both groups; and (3) items with low correct response rates in both groups. Since no prior validation or reference data were available for this knowledge test, a ≥40% difference in correct response rates was defined on an ad hoc basis as a clinically meaningful threshold. This cutoff was informed by the observed between-group separation in overall knowledge scores and was intended to identify items with substantial practical relevance rather than minor statistical differences.

#### Quality of Life

Quality of life was assessed using the Japanese version of the Kansas City Cardiomyopathy Questionnaire Short version (KCCQ-12).^7,11,12^ The KCCQ-12 is a disease-specific, self-administered instrument consisting of 23 items covering physical limitations, symptom frequency and severity, symptom stability, self-efficacy, social limitations, and quality of life. Each domain is scored from 0 to 100, with higher scores indicating better health status. Domain scores are averaged to generate an overall summary score. A change of 5 points or more in the KCCQ-12 Score is generally considered a small but clinically meaningful improvement, whereas changes of 10 and 20 points represent moderate and large improvements, respectively.

#### Clinical outcomes

Clinical outcomes included all-cause mortality and HF rehospitalization within one year after discharge. Events were identified from hospital records chart reivew, or confirmed by telephone or postal inquiries to patients, caregivers, treating physicians, or care facilities if outpatient follow-up was interrupted. All events were adjudicated by more than two invetigators to ensure consistency for HF hospitalization.

#### Statistical Analysis

All analyses were conducted using a conservative approach, assuming non-normal distributions for continuous variables and accounting for the small sample size in categorical analyses. Accordingly, continuous variables are presented as medians with interquartile ranges, and categorical variables as counts and percentages. Between-group comparisons were performed using the Wilcoxon rank-sum test for continuous variables and Fisher’s exact test for categorical variables, as appropriate. Between-group differences at each time point were evaluated using the Wilcoxon rank-sum test. No adjustment was made for multiple comparisons.

Qualitative data from semi-structured interviews were analyzed inductively using thematic analysis. Audio-recorded interviews were transcribed verbatim and coded independently by two investigators without the use of a predefined coding framework. Codes were iteratively grouped into categories based on conceptual similarity, and emerging themes were discussed among investigators until consensus was achieved. Representative quotations were selected to illustrate key themes and enhance interpretive transparency.

All statistical analyses were performed using Stata/SE version 15 (StataCorp LLC, College Station, TX, USA) for Mac, and a two sided p value <0.05 was considered statistically significant.

## Results

### Characteristics during index HF hospitalization

Between April and December 2021, 176 patients were hospitalized for acute heart failure under the care of five cardiologists (**Figure 1**). Of these, 101 were excluded due to cognitive impairment or insufficient capacity for self-care, as determined by consensus of at least two members of the investigator team. The remaining 75 patients were invited to participate in the knowledge assessment, symptom questionnaire, and follow-up evaluations. Nineteen patients were subsequently excluded (12 declined participation and 7 did not meet eligibility criteria). Ultimately, 56 patients completed all assessments and comprised the analytical cohort. Among them, 31 were classified as the knowledge-improved group, while 25 were classified as the non-improved group.

**Figure x1.**
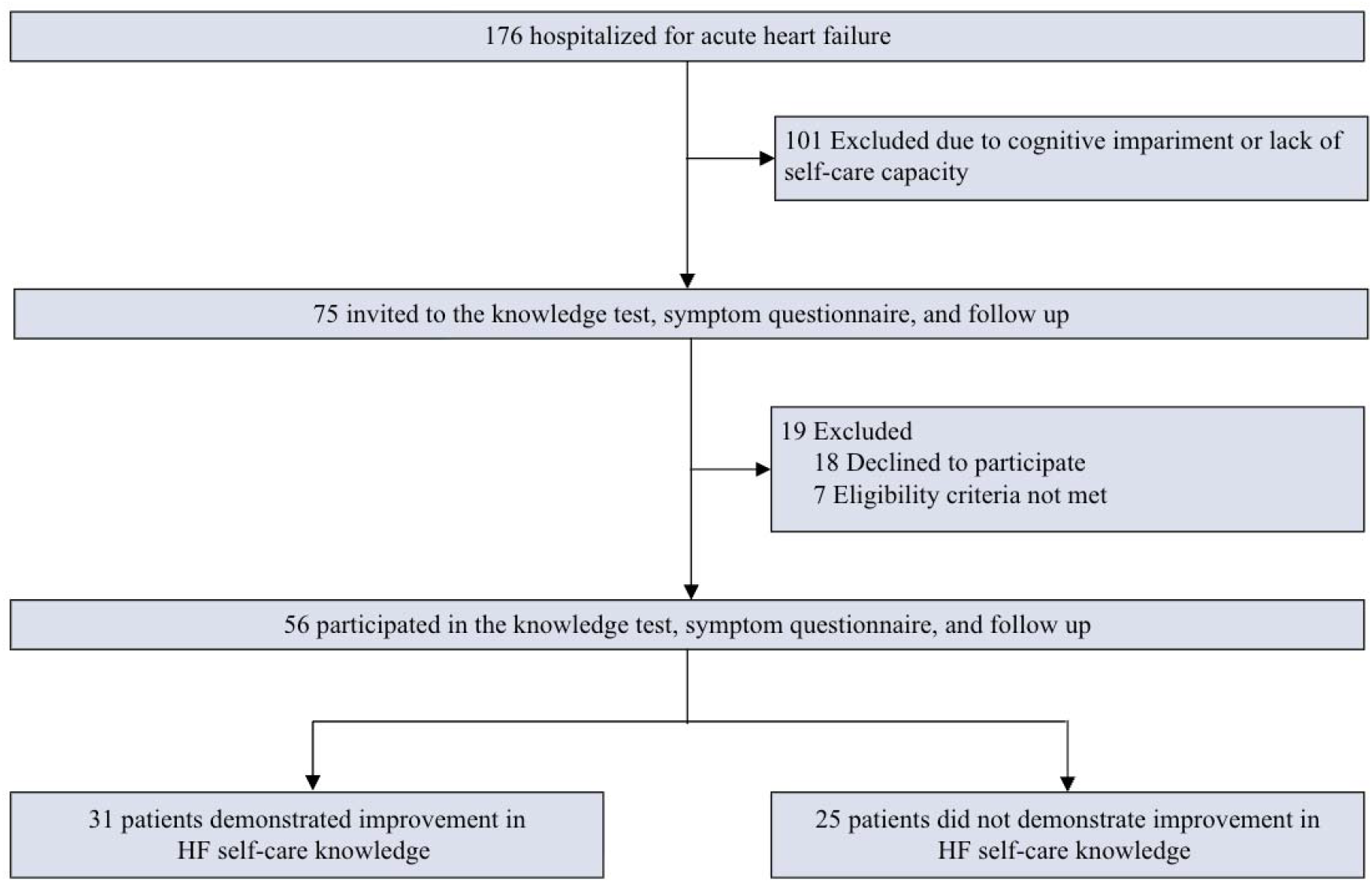
Flow diagram of patient selection and participation. A total of 176 patients were hospitalized for acute heart failure. Of these, 101 were excluded due to significant cognitive impairment or lack of self-care capacity. The remaining 75 patients were invited to participate in the knowledge test, symptom questionnaire, and follow-up. Nineteen patients were further excluded (18 declined to participate, 7 did not meet eligibility criteria). Ultimately, 56 patients completed all assessments. Among them, 31 patients demonstrated improvement in heart failure HF self-care knowledge, whereas 25 patients did not show such improvement. Abbreviations: HF = Heart Failure.

Upon admission, patients had a median age of 78 years (IQR, 66–84), and 66% were men. Nearly half (46%) had heart failure with reduced ejection fraction, with a median left ventricular ejection fraction of 41% (IQR, 29–53) (**Table 1**). Overall disease severity was moderate, reflected by a median MAGGIC risk score of 26 (IQR, 22–29). Admission demographics, comorbidities, vital signs, heart failure phenotype, prior heart failure hospitalization, and laboratory findings—including B-type natriuretic peptide—were well balanced between the knowledge-improved and non-improved groups.

**Table 1.**
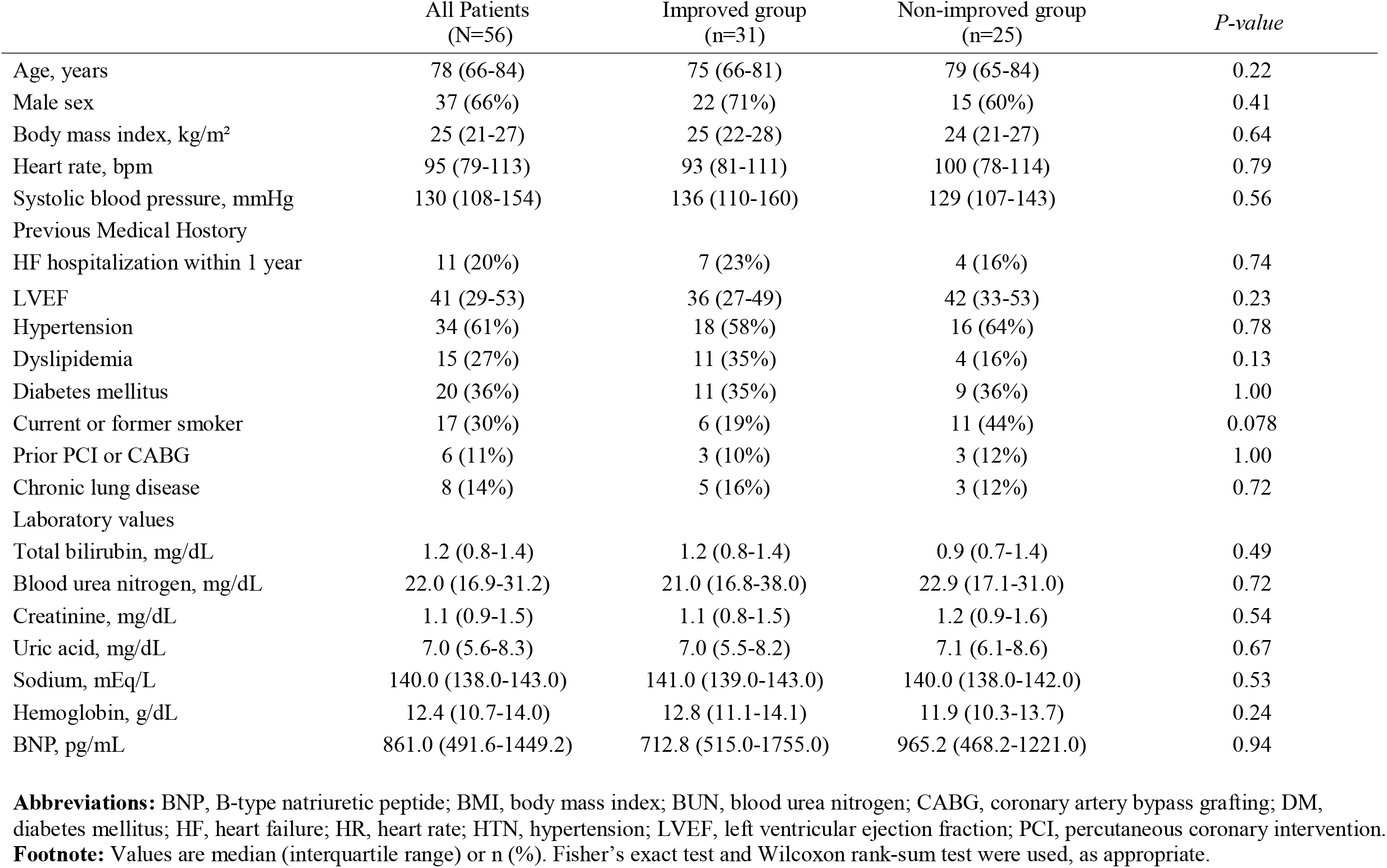
Patient Characteristics at Admission.

By the time of discharge, clinical profiles remained largely comparable between groups, including hemodynamics, body weight and in-hospital weight change, renal function, serum sodium, hemoglobin, B-type natriuretic peptide levels, MAGGIC risk score, length of stay, discharge destination, and use of guideline-directed medical therapies **(Table 2)**. In contrast, markers related to nutritional status differed between groups. The knowledge-improved group had higher serum albumin levels at discharge (median 3.8 [IQR, 3.5–4.2] vs. 3.4 [3.1–3.7] g/dL; *P* = 0.03) and demonstrated better dietary intake during the final days of hospitalization, with higher consumption of both main dishes (median score 10 [10–10] vs. 10 [7–10]; *P* = 0.01) and side dishes (10 [10–10] vs. 10 [8–10]; *P* = 0.02) compared with the non-improved group. KCCQ overall summary scores were similar between groups before discharge (median score 46 [37–63] vs. 39 [29–57] ; *P* = 0.38).

**Table 2.**
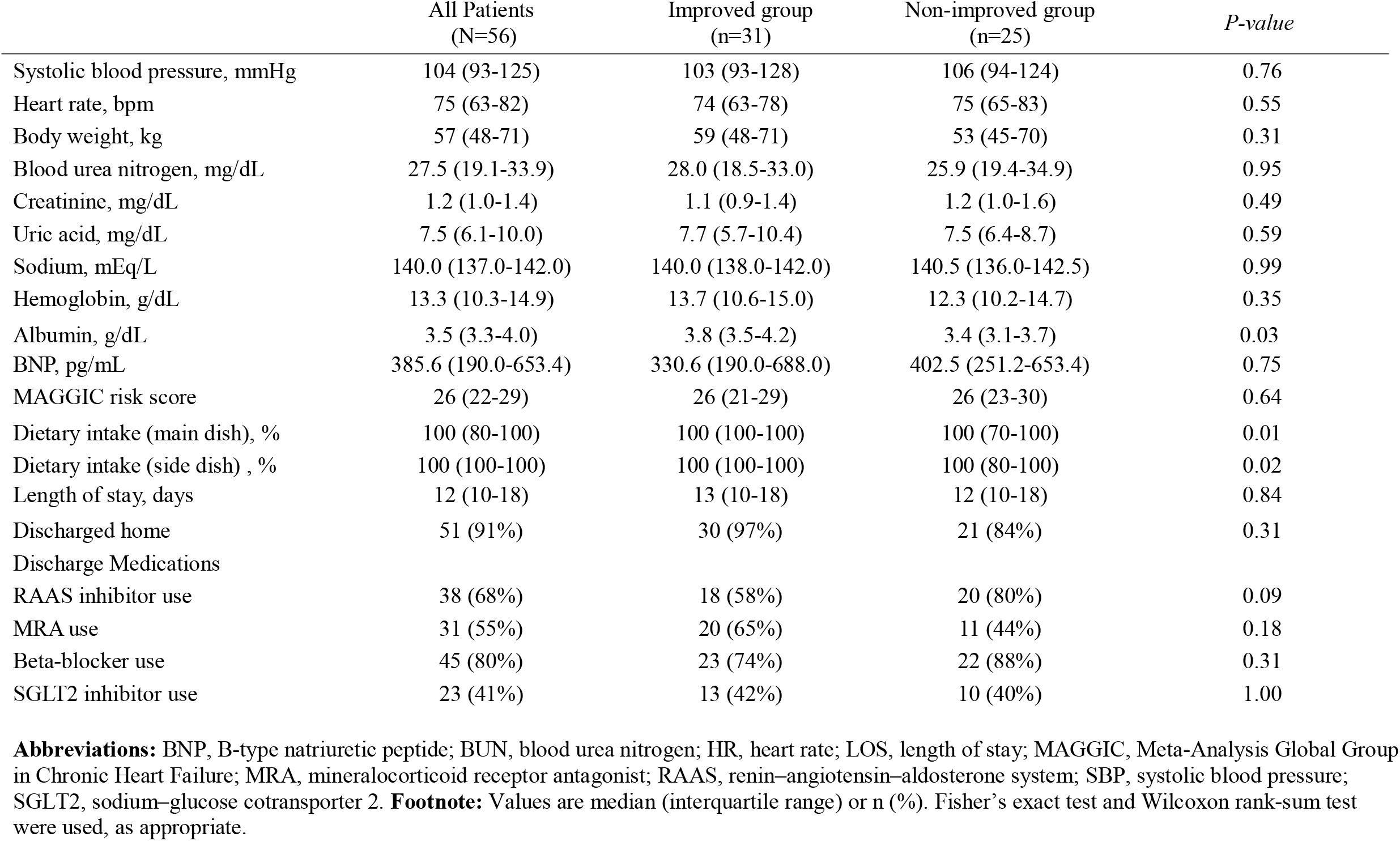
Patient Characteristics at Discharge.

|  | All Patients<br>(N=56) | Improved group<br>(n=31) | Non-improved group<br>(n=25) | *P-value* |
|---|---|---|---|---|
| Systolic blood pressure, mmHg | 104 (93-125) | 103 (93-128) | 106 (94-124) | 0.76 |
| Heart rate, bpm | 75 (63-82) | 74 (63-78) | 75 (65-83) | 0.55 |
| Body weight, kg | 57 (48-71) | 59 (48-71) | 53 (45-70) | 0.31 |
| Blood urea nitrogen, mg/dL | 27.5 (19.1-33.9) | 28.0 (18.5-33.0) | 25.9 (19.4-34.9) | 0.95 |
| Creatinine, mg/dL | 1.2 (1.0-1.4) | 1.1 (0.9-1.4) | 1.2 (1.0-1.6) | 0.49 |
| Uric acid, mg/dL | 7.5 (6.1-10.0) | 7.7 (5.7-10.4) | 7.5 (6.4-8.7) | 0.59 |
| Sodium, mEq/L | 140.0 (137.0-142.0) | 140.0 (138.0-142.0) | 140.5 (136.0-142.5) | 0.99 |
| Hemoglobin, g/dL | 13.3 (10.3-14.9) | 13.7 (10.6-15.0) | 12.3 (10.2-14.7) | 0.35 |
| Albumin, g/dL | 3.5 (3.3-4.0) | 3.8 (3.5-4.2) | 3.4 (3.1-3.7) | 0.03 |
| BNP, pg/mL | 385.6 (190.0-653.4) | 330.6 (190.0-688.0) | 402.5 (251.2-653.4) | 0.75 |
| MAGGIC risk score | 26 (22-29) | 26 (21-29) | 26 (23-30) | 0.64 |
| Dietary intake (main dish), % | 100 (80-100) | 100 (100-100) | 100 (70-100) | 0.01 |
| Dietary intake (side dish) , % | 100 (100-100) | 100 (100-100) | 100 (80-100) | 0.02 |
| Length of stay, days | 12 (10-18) | 13 (10-18) | 12 (10-18) | 0.84 |
| Discharged home | 51 (91%) | 30 (97%) | 21 (84%) | 0.31 |
| Discharge Medications |  |  |  |  |
| RAAS inhibitor use | 38 (68%) | 18 (58%) | 20 (80%) | 0.09 |
| MRA use | 31 (55%) | 20 (65%) | 11 (44%) | 0.18 |
| Beta-blocker use | 45 (80%) | 23 (74%) | 22 (88%) | 0.31 |
| SGLT2 inhibitor use | 23 (41%) | 13 (42%) | 10 (40%) | 1.00 |

**Abbreviations:** BNP, B-type natriuretic peptide; BUN, blood urea nitrogen; HR, heart rate; LOS, length of stay; MAGGIC, Meta-Analysis Global Group in Chronic Heart Failure; MRA, mineralocorticoid receptor antagonist; RAAS, renin–angiotensin–aldosterone system; SBP, systolic blood pressure; SGLT2, sodium–glucose cotransporter 2. **Footnote:** Values are median (interquartile range) or n (%). Fisher’s exact test and Wilcoxon rank-sum test were used, as appropriate.

**Table 3.** Changes in Heart Failure Knowledge and KCCQ Scores at Pre-education, Post-education, and Initial Outpatient Follow-up.

|  | All Patients<br>(N=56) | Improved group<br>(n=31) | *P-value* | Non-improved group<br>(n=25) | *P-value* |
|---|---|---|---|---|---|
| Knowledge test, score |  |  |  |  |  |
| Pre-education | 10 (7-13) | 10 (6-13) |  | 10 (8-13) |  |
| Post-education | 15 (12-18) | 17 (14-19) | <0.001 | 12 (9-14) | <0.001 |
| Initial outpatient visit | 15 (11-17) | 16 (12-18) | <0.001 | 13 (10-16) | <0.001 |
| KCCQ-12 Score |  |  |  |  |  |
| Post-education | 45 (33-58) | 46 (37-63) |  | 39 (29-57) |  |
| Initial outpatient visit | 63 (46-83) | 63 (48-85) | 0.008 | 62 (45-80) | 0.14 |

**Abbreviations:** KCCQ, Kansas City Cardiomyopathy Questionnaire; OSS, Overall Summary Score; HF, heart failure. **Footnote:** Values are median (interquartile range). P values shown for post-education and initial outpatient visit indicate within-group changes from pre-education, evaluated using the Wilcoxon signed-rank test. Between-group differences at each time point were evaluated using the Wilcoxon rank-sum test. No adjustment was made for multiple comparisons.

**Table 4.** Item-Level Differences in Correct Response Rates Following Nurse-Led Heart Failure Education.

| Questions | Improved group (n=31) | Non-improved group (n=25) | % Difference | *P-value* |
|---|---|---|---|---|
| Q8: It is better to avoid exercise as much as possible. | 26 (83.9%) | 8 (40.0%) | 43.9% | 0.001 |
| Q7: Worsening heart failure can cause more frequent urination. | 29 (93.5%) | 10 (50.0%) | 43.5% | <0.001 |
| Q10: The recommended daily salt intake is less than 10 g per day. | 24 (77.4%) | 7 (35.0%) | 42.4% | 0.002 |
| Q14: Even a cold or mild diarrhea can worsen heart failure. | 22 (73.3%) | 6 (31.6%) | 41.7% | 0.004 |
| Q18: Municipalities keep an emergency medical information card at home entrances to share patients’ wishes during emergency transport. | 13 (41.9%) | 2 (10.5%) | 31.4% | 0.02 |
| Q16: Writing down instructions in advance about medical decisions if you lose decision making capacity is called a living will. | 24 (77.4%) | 9 (47.4%) | 30.0% | 0.03 |
| Q17: Decisions stated in a living will cannot be changed later. | 24 (77.4%) | 9 (47.4%) | 30.0% | 0.03 |
| Q13: Excessive fluid intake can worsen heart failure. | 21 (67.7%) | 8 (40.0%) | 27.7% | 0.051 |
| Q19: Poor oral hygiene can cause gum inflammation and increase the risk of atherosclerosis and bacteria entering the heart. | 30 (96.8%) | 14 (73.7%) | 23.1% | 0.02 |
| Q15: Thinking about and discussing what matters to you and your preferences for medical care is called advance care planning. | 28 (90.3%) | 13 (68.4%) | 21.9% | 0.05 |
| Q9: Exercise should be stopped if shortness of breath or chest pain occurs. | 26 (83.9%) | 13 (65.0%) | 18.9% | 0.12 |
| Q1: If symptoms improve, heart failure can be considered completely cured. | 28 (90.3%) | 15 (75.0%) | 15.3% | 0.14 |
| Q3: Heart failure is classified into three stages. | 12 (40.0%) | 5 (25.0%) | 15.0% | 0.27 |
| Q20: As a rule, blood thinning medications should be continued during dental treatment. | 16 (51.6%) | 7 (36.8%) | 14.8% | 0.31 |
| Q12: One bowl of miso soup (180 mL) contains about 0.5 g of salt. | 29 (93.5%) | 16 (80.0%) | 13.5% | 0.14 |
| Q2: Symptoms of heart failure include shortness of breath, difficulty breathing when lying flat, and leg swelling. | 30 (96.8%) | 17 (85.0%) | 11.8% | 0.13 |
| Q11: One pickled plum (10 g) contains about 0.5 g of salt. | 29 (93.5%) | 16 (84.2%) | 9.3% | 0.29 |
| Q5: Using unit dose packaging or a medication calendar is effective in preventing missed doses. | 31 (100.0%) | 19 (95.0%) | 5.0% | 0.21 |
| Q4: If you feel better, it is acceptable to stop taking your medication. | 29 (93.5%) | 18 (90.0%) | 3.5% | 0.65 |
| Q6: Carrying a heart failure handbook and a medication notebook helps healthcare providers understand your care during visits. | 31 (100%) | 20 (100%) | 0.0% | NA |

**Abbreviations:** HF, heart failure. **Footnotes:** Values are n (%). All questions were answered as *Yes / No / I do not know*. Between-group differences were calculated as absolute percentage differences. P values were derived using Fisher’s exact test. *NA* indicates not applicable due to identical response rates between groups.

### Heart failure knowledge at pre, post education, and initial outpatient visit

Heart failure knowledge test scores at baseline, prior to nurse-led education, were similar between groups (median [IQR]: 10 [6–13] in the improved group and 10 [8–13] in the non-improved group; *P* = 0.95). Following nurse-led discharge education, knowledge scores increased in both groups but differed in magnitude, with higher post-education scores observed in the improved group (17 [14–19]) compared with the non-improved group (12 [9–14]). Similarly, at the initial outpatient visit, knowledge scores remained higher in the improved group (16 [12–18]) than in the non-improved group (13 [10–16]; *P* = 0.021). Within-group comparisons using paired Wilcoxon signed-rank tests demonstrated significant increases in knowledge scores from baseline to post-education and to the outpatient visit in both groups.

Item level analysis identified four questions with absolute differences greater than 40% in correct response rates between groups, indicating substantial knowledge separation after nurse led heart failure education (**Table 2**). The largest difference was observed for the item “It is better to avoid exercise as much as possible” (Q8), which was correctly answered by 83.9% (26/31) of patients in the knowledge improved group compared with 40.0% (8/25) in the non improved group (difference 43.9%, P = 0.001). Similarly, the item “Heart failure may cause frequent urination at night” (Q7) showed correct response rates of 93.5% (29/31) versus 50.0% (10/25), respectively (difference 43.5%, P < 0.001). A large difference was also observed for the question “One pickled plum (10 g) contains 0.5 g of salt” (Q10), with correct responses in 77.4% (24/31) of the knowledge improved group and 35.0% (7/25) of the non improved group (difference 42.4%, P = 0.002). In addition, for the item “Even mild diarrhea can worsen heart failure” (Q14), correct response rates were 73.3% (22/31) in the knowledge improved group and 31.6% (6/25) in the non improved group (difference 41.7%, P = 0.004).

### Post-Discharge Outcomes

Days from discharge to the initial outpatient visit were 14 [11–21] days overall, 14 [11–18] days in the knowledge-improved group, and 12 [11–24] days in the non-improved group, with no significant difference between groups. Heart failure handbook diary completion for recording vital signs, body weight, and symptoms was similar across groups (41 [73%] overall; 23 [74%] in the improved group; 18 [72%] in the non-improved group; *P* = 0.29). Of note, KCCQ-12 Score data at the initial outpatient visit were available for 38 of 48 patients and were similar between groups (63 [48–85] vs. 62 [45–80]). The change in KCCQ-12 Score from pre-education to the initial outpatient visit was similar between groups (median [IQR]: 19 [−5 to 42] in the improved group vs. 14 [−6 to 41] in the non-improved group; *P* = 0.86). The proportion of patients achieving a clinically meaningful improvement in KCCQ-12 Score(>5-point increase) was similar between groups (overall: 21 [64%]; improved group: 13 [68%]; non-improved group: 8 [57%]; P = 0.72).

During the one-year follow-up period, 5 patients (8.9%) were lost to follow-up, resulting in a follow-up completion rate of 91.1%. Among patients with available follow-up data, no significant differences in clinical outcomes were observed between groups. All-cause mortality occurred in 3 of 28 patients (10.7%) in the knowledge-improved group and in 1 of 23 patients (4.3%) in the non-improved group (P = 0.40). Heart failure–related readmission occurred in 5 of 28 patients (17.9%) in the knowledge-improved group and in 3 of 23 patients (13.0%) in the non-improved group (P = 0.64).

### Qualitative Analysis

A total of four patients, three in the knowledge improved group and one in the non improved group, who were readmitted during the study period underwent semi structured interviews, from which 94 codes were identified and organized into 26 subcategories and 9 overarching categories. These categories captured patients’ experiences related to understanding the handbook content and recording daily vital signs and symptoms. They included: (1) recording daily information in the handbook using personalized strategies; (2) using handbook records to support disease management and lifestyle adjustment; (3) incorporating knowledge gained through education into daily self care; (4) experiencing challenges in disease management despite regular handbook use; (5) feeling connected to healthcare professionals through the handbook; (6) accepting heart failure through continued engagement with the handbook; (7) reflecting on future care and end of life preferences; (8) perceiving inconvenience related to the handbook content or format; and (9) wishing that heart failure education had been provided earlier, before disease worsening.

## Discussion

In this prospective mixed-methods registry, we observed several associations related to nurse-led heart failure education in routine clinical practice. First, heart failure knowledge levels varied across patients over time, with persistent differences between those with higher and lower knowledge scores, particularly in domains related to exercise, salt intake, symptom recognition, and exacerbating factors. Second, higher knowledge scores were associated with more favorable markers of nutritional status during hospitalization, including higher serum albumin levels and more complete dietary intake, despite similar baseline characteristics, in-hospital management, and rates of guideline-directed medical therapy at discharge. Third, qualitative findings complemented these observations by illustrating that patients commonly encountered challenges in integrating knowledge into long-term self-care within the context of disease progression and psychosocial constraints.

Our study aligns with and adds to the existing literature by clarifying specific areas in which patients may have difficulty understanding foundational knowledge required to live with heart failure as a chronic condition. The importance of patient education is emphasized in the Japanese heart failure guidelines, and both the American College of Cardiology and the American Heart Association recommend that all patients with symptomatic heart failure, irrespective of ejection fraction, receive structured education focused on self-care.^3,13^ However, most prior clinical trials in this field have been conducted in Western countries, with relatively few studies from the Asia–Pacific region, where the average age of patients with heart failure is higher, raising concerns about the generalizability of these findings.^1^ Moreover, many interventions evaluated in previous trials consisted of intensive, multi-component nurse-led programs that combined predischarge education with frequent follow-up contacts, which may not fully reflect routine, feasible clinical practice.^6,14–16^ In this context, predischarge education remains a central component of real-world heart failure care. In our study, heart failure knowledge levels varied across patients over time, with persistent differences observed between those with higher and lower overall knowledge scores. These differences were most evident in domains related to exercise, salt intake, symptom recognition, and recognition of exacerbating factors. Item-level analyses highlighted gaps in understanding regarding physical activity (“It is better to avoid exercise as much as possible”), symptom awareness (“Heart failure may cause frequent urination at night”), dietary salt content (“One pickled plum [umeboshi 10 g] contains 0.5 g of salt”), and triggers of worsening (“Even mild diarrhea can worsen heart failure”). Together, these domains span multiple aspects of daily self-care and highlight that knowledge represents only part of the journey, with translation into everyday self-management remaining an additional challenge after discharge.

Second, higher knowledge scores were associated with more favorable markers of nutritional status during hospitalization, including higher serum albumin levels and more complete dietary intake, despite similar baseline characteristics, in-hospital management, and rates of guideline-directed medical therapy at discharge. Cognitive function and nutritional status have been reported to be closely associated in patients with heart failure, particularly among older adults, where they may reflect overall disease severity, the effects of decongestion, or the presence of cachexia.^17–19^ Although standardized measures of nutritional status and frailty, such as the Geriatric Nutritional Risk Index or the Clinical Frailty Scale, were not assessed in this study, the observed tendency toward lower intake of energy via protein-rich side dishes warrants consideration.^20–23^ Reduced protein intake may be associated with loss of skeletal muscle mass, which in turn is linked to diminished physical function and lower levels of daily activity.^24^ Such interpretations remain speculative in the context of the present study and should not be overstated. Nonetheless, emerging evidence highlights the potential importance of preserving or restoring muscle mass and physical function during hospitalization and the post-discharge period, including through structured rehabilitation programs, as part of comprehensive care for patients with heart failure.^25^

Third, the qualitative findings added contextual depth by showing that patients who experienced heart failure worsening and subsequent readmission often struggled to translate knowledge into sustained self-care amid disease progression and psychosocial constraints. Notably, three of the four interviewed patients were classified in the knowledge-improved group, suggesting that higher knowledge levels alone may not be sufficient in the setting of more advanced disease that may not be sufficiently reflected via risk prediction modeling. Of note, the KCCQ-12 scores prior to discharge was comparable to those observed in the China PEACE 5p-HF (China Patient-Centered Evaluative Assessment of Cardiac Events Prospective Heart Failure) study where patients were asked to answer the KCCQ-12 within 48 hour of index admission for acute heart failure.^26^ These readmitted patients exhibited features consistent with more severe heart failure phenotypes, such as dilated cardiomyopathy at younger ages, in which ongoing clinical deterioration may constrain the practical impact of education alone. In such situations, effective management may require more individualized and layered support involving family members, nurses, and other caregivers, extending beyond what can be achieved through a single educational resource. These observations are consistent with prior qualitative studies that emphasize the importance of tailored educational approaches for people living with heart failure.^27^ While structured educational materials and periodic reinforcement may remain valuable, these findings underscore the limitations of self-care strategies based solely on knowledge and highlight the importance of timely clinical evaluation when symptoms worsen.

## Limitations

Several limitations should be acknowledged, alongside important strengths. This was a single center observational study conducted at one institution in a Japanese metropolitan area setting, which may limit generalizability to other health care systems and patient populations. The heart failure handbook and the associated knowledge assessment were developed locally and pragmatically to align with routine practice and were not externally validated, which may affect applicability to other settings, although this design reflects how education is commonly delivered in daily care. The definition of knowledge improved and non improved groups was determined on an ad hoc basis and was not predefined according to score distributions or external benchmarks, but repeated assessments allowed evaluation of knowledge patterns over time rather than reliance on a single measurement. The overall sample size was modest, particularly for the qualitative substudy, limiting statistical power to detect differences in clinical outcomes, yet the mixed methods approach provided complementary quantitative and experiential insights. Potentially relevant factors such as baseline cognitive function, caregiver involvement, socioeconomic status, and health literacy were not systematically or quantitavely assessed, raising the possibility of unmeasured confounding; nonetheless, inclusion required agreement by at least two investigators regarding cognitive capacity, supporting consistency in patient selection. The study was conducted during the COVID-19 pandemic, when health care utilization and patient behavior were atypical and may have influenced educational delivery and outcomes. Finally, the one year follow up period may not capture longer term associations between patient education, self care, and clinical outcomes, although the prospective design and early post discharge assessments provide valuable information on short term trajectories.

## Conclusion

In this single-center prospective registry, structured heart failure education was associated with differences in knowledge levels, while no significant differences were observed in quality of life, one-year mortality, or readmission rates. Quantitative and qualitative findings together suggest that heart failure education relates to both cognitive knowledge and the social and experiential aspects of self-care. These results indicate that educational approaches may need to be tailored to individual patient profiles, including disease severity and progression, as well as cognitive, educational, and physical function.

## Data Availability

The datasets generated and analyzed during the current study are not publicly available due to patient privacy considerations but are available from the corresponding author on reasonable request.

