## Supplemental Materials for "Nurse-Led Heart Failure Education and Patient Outcomes: A Mixed-Methods Prospective Registry Study"

Supplemental Material

Supplemental Table 1. Heart Failure Knowledge Questionnaire: Original Items and English Translation

Supplemental Table 1. Heart Failure Knowledge Questionnaire: Original Items and English Translation

| **Question No.** | **Japanese (Original)** | **English Translation** |
| --- | --- | --- |
| Q1 | 苦しい症状がよくなれば、心不全は完全になおったと考えてよい。 | If symptoms improve, heart failure can be considered completely cured. |
| Q2 | 心不全の症状には、「息苦しさ」「横になると呼吸が苦しい」「足がむくむ」などがある。 | Symptoms of heart failure include shortness of breath, difficulty breathing when lying flat, and leg swelling. |
| Q3 | 心不全には3つのステージがあるとされている。 | Heart failure is classified into three stages. |
| Q4 | 体調がよくなったと感じたら、薬をやめてもよい。 | If you feel better, it is acceptable to stop taking your medication. |
| Q5 | 薬の飲み忘れを防ぐには、お薬の一包化やお薬カレンダーを用いることが有効である。 | Using unit-dose packaging or a medication calendar is effective in preventing missed doses. |
| Q6 | 「心不全手帳」と「お薬手帳」をいつも持ち歩くことで、受診の際、医療従事者が診療内容を把握しやすい。 | Carrying a heart failure handbook and a medication notebook helps healthcare providers understand your care during visits. |
| Q7 | 心不全の悪化でトイレが近くなることがある。 | Worsening heart failure can cause more frequent urination. |
| Q8 | なるべく運動をしない方がよい。 | It is better to avoid exercise as much as possible. |
| Q9 | 息切れや胸の痛みが出れば運動は中止した方がよい。 | Exercise should be stopped if shortness of breath or chest pain occurs. |
| Q10 | 食事の塩分摂取量の目安は、1日10g未満である。 | The recommended daily salt intake is less than 10 g per day. |
| Q11 | 梅干し1粒（10g）は塩分約0.5gである。 | One pickled plum (10 g) contains about 0.5 g of salt. |
| Q12 | 味噌汁1杯（180 mL）は塩分約0.5gである。 | One bowl of miso soup (180 mL) contains about 0.5 g of salt. |
| Q13 | 水分をとりすぎて心不全が悪くなることがある。 | Excessive fluid intake can worsen heart failure. |
| Q14 | 風邪やちょっとした下痢でも、心不全が悪くなることがある。 | Even a cold or mild diarrhea can worsen heart failure. |
| Q15 | 万が一の時に備えて、自分の大切にしていることや、どのような医療やケアを望んでいるかを考え、話し合ったりすることを、アドバンス・ケア・プランニング（これからの医療やケアに関する人生会議）という。 | Thinking about and discussing what matters to you and your preferences for medical care is called advance care planning. |
| Q16 | 自分が病気や事故で意思能力の回復が見込めない状態になった場合、自分自身が受ける医療の決定に関する指示をあらかじめ書面等に記しておくことを「リビングウィル」という。 | Writing down instructions in advance about medical decisions if you lose decision-making capacity is called a living will. |
| Q17 | 「リビングウィル」で意思表示したことは、最期まで変更することができない。 | Decisions stated in a living will cannot be changed later. |
| Q18 | 市町村では救急搬送時に在宅医療機関のスタッフが、医療情報に関する本人の意思を共有する目的で「救急医療情報カード」を、玄関に大切に保管している。 | Municipalities keep an emergency medical information card at home entrances to share patients’ wishes during emergency transport. |
| Q19 | 口の中が汚くなると、歯肉の炎症を起こし、動脈硬化の進行や心臓の中に細菌が入りやすくなる。 | Poor oral hygiene can cause gum inflammation and increase the risk of atherosclerosis and bacteria entering the heart. |
| Q20 | 歯の治療を受ける際は原則として、血をサラサラにする薬は継続したまま行う。 | As a rule, blood-thinning medications should be continued during dental treatment. |

Supplemental Table 2. Interview Questions
